# Hydroxychloroquine as pre-exposure prophylaxis for COVID-19 in healthcare workers: a randomized trial

**DOI:** 10.1101/2020.09.18.20197327

**Authors:** Radha Rajasingham, Ananta S Bangdiwala, Melanie R Nicol, Caleb P Skipper, Katelyn A Pastick, Margaret L Axelrod, Matthew F Pullen, Alanna A Nascene, Darlisha A Williams, Nicole W Engen, Elizabeth C Okafor, Brian I Rini, Ingrid A Mayer, Emily G McDonald, Todd C. Lee, Peter Li, Lauren J MacKenzie, Justin M Balko, Stephen J Dunlop, Katherine H Hullsiek, David R Boulware, Sarah M Lofgren, on behalf of the COVID PREP team

## Abstract

**Background:** Severe acute respiratory syndrome coronavirus 2 (SARS-CoV-2) is a rapidly emerging virus causing the ongoing Covid-19 pandemic with no known effective prophylaxis. We investigated whether hydroxychloroquine could prevent SARS CoV-2 in healthcare workers at high-risk of exposure.

**Methods:** We conducted a randomized, double-blind, placebo-controlled clinical trial of healthcare workers with ongoing exposure to persons with Covid-19, including those working in emergency departments, intensive care units, Covid-19 hospital wards, and first responders.

Participants across the United States and in the Canadian province of Manitoba were randomized to hydroxychloroquine 400mg once weekly or twice weekly for 12 weeks. The primary endpoint was confirmed or probable Covid-19-compatible illness. We measured hydroxychloroquine whole blood concentrations.

**Results:** We enrolled 1483 healthcare workers, of which 79% reported performing aerosol-generating procedures. The incidence of Covid-19 (laboratory-confirmed or symptomatic compatible illness) was 0.27 events per person-year with once-weekly and 0.28 events per person-year with twice-weekly hydroxychloroquine compared with 0.38 events per person-year with placebo. For once weekly hydroxychloroquine prophylaxis, the hazard ratio was 0.72 (95%CI 0.44 to 1.16; P=0.18) and for twice weekly was 0.74 (95%CI 0.46 to 1.19; P=0.22) as compared with placebo. Median hydroxychloroquine concentrations in whole blood were 98 ng/mL (IQR, 82-120) with once-weekly and 200 ng/mL (IQR, 159-258) with twice-weekly dosing. Hydroxychloroquine concentrations did not differ between participants who developed Covid-19 (154 ng/mL) versus participants without Covid-19 (133 ng/mL; P=0.08).

**Conclusions:** Pre-exposure prophylaxis with hydroxychloroquine once or twice weekly did not significantly reduce laboratory-confirmed Covid-19 or Covid-19-compatible illness among healthcare workers.

**Key Points:** In this randomized clinical trial of 1483 high-risk healthcare workers, there was no significant reduction in incidence of Covid-19 with once weekly or twice weekly hydroxychloroquine compared to placebo.

## Background

Covid-19 creates a substantial strain on the healthcare system with frontline healthcare workers at increased risk of infection, and yet they are simultaneously essential for sustaining an adequate emergency response. Unfortunately, at present, no effective oral chemoprophylaxis or vaccination against Covid-19 exists. On August 4, 2020, the Centers for Disease Control and Prevention (CDC) reported over 121,000 cases of Covid-19 among healthcare personnel in the United States.(1) An effective pre-exposure prophylaxis medication for healthcare workers with repeated SARS CoV-2 exposure, even if only partially effective, would be a powerful public health tool to reduce transmission of SARS-CoV-2 and protect frontline workers from Covid-19.

While intensive efforts are being directed towards treatment discovery and vaccine development, repurposing existing medications is a more swift and economical approach to fulfill a time-sensitive need for effective prophylaxis. Chloroquine has demonstrated *in vitro* activity against SARS-CoV and SARS-CoV-2.(2, 3) Recent studies demonstrated that hydroxychloroquine, a derivative molecule of chloroquine, is also active against SARS-CoV-2 and may demonstrate greater *in vitro* viral inhibition.(4, 5) However, it remains unclear if *in vitro* activity corresponds to clinical efficacy. Randomized clinical trials in post-exposure prophylaxis, early outpatient treatment, and inpatient treatment have not borne out this initial promise.(6-9) Nonetheless, some have postulated that the post-exposure and early treatment trials may not have achieved therapeutic concentrations early enough to have demonstrated a benefit (6). In India, hydroxychloroquine 400mg weekly is recommended nationally in asymptomatic healthcare workers at high risk for Covid-19, despite no substantial evidence that it prevents Covid-19.(10)

There is ongoing interest in the concept of pre-exposure prophylaxis whereby a patient has already achieved adequate drug concentrations at the time of viral exposure. Therefore, we sought to determine the effectiveness of hydroxychloroquine as pre-exposure prophylaxis in healthcare workers at high-risk of SARS-CoV-2 exposure in a randomized, placebo-controlled clinical trial setting.

## Methods

### Study Design

We conducted a randomized, double-blind, placebo-controlled clinical trial (Clinicaltrials.gov NCT04328467) to evaluate whether hydroxychloroquine could prevent Covid-19 in high-risk healthcare workers across the United States and Canada. Enrollment began on April 6, 2020, and ended May 26, 2020; follow up was completed on July 13, 2020.

Participants were randomly assigned in a 2:2:1:1 ratio to receive hydroxychloroquine given as a loading dose of 400mg (two 200mg tablets) twice separated by 6-8 hours followed by (i) 400 mg (two 200mg tablets) <u>once</u> weekly for 12 weeks or (ii) 400 mg (two 200mg tablets) <u>twice</u> weekly for 12 weeks, or to placebo which was prescribed in a matched fashion including a loading dose of two tablets followed by two tablets once or twice weekly for 12 weeks.

### Participants

We included healthcare workers aged 18 years and older with ongoing exposure to persons with Covid-19. A high-risk healthcare worker was defined as working in an emergency department or intensive care unit, on a dedicated Covid-19 hospital ward, as a first responder, or whose job description included regularly performing aerosol-generating procedures (e.g., anesthesiologists or otolaryngologists), and included physicians, nurses, advanced practice providers, and other personnel (e.g., respiratory therapists).

We excluded persons who reported active or prior Covid-19 (either confirmed or symptom-compatible illness), no expected exposure to patients, or contraindication to hydroxychloroquine (**Appendix**).

### Setting

We enrolled participants nationwide in the United States and the Canadian province of Manitoba. We recruited participants using social media platforms targeting healthcare providers. Participants self-enrolled via a secure internet-based survey using the Research Electronic Data Capture (REDCap) system.(11) Participants provided a digitally captured informed consent signature after passing a comprehension assessment.

### Study Assessments

Online study assessments were scheduled at enrollment, medication initiation, and weekly after enrollment. Each assessment included a report of study medication adherence, medication side effects, the number of patient-facing contact hours, contact with patients with confirmed or possible Covid-19, personal protective equipment (PPE) use, Covid-19 compatible symptoms, SARS-CoV-2 testing results, and any hospitalization.

### Outcomes

The primary outcome was Covid-19–free survival time by laboratory-confirmed or probable compatible illness. Confirmed Covid-19 was defined as SARS-CoV-2 polymerase chain reaction (PCR) positivity by self-report. Given limited availability of outpatient PCR testing in many jurisdictions during our study period, particularly in April 2020, probable Covid-19 based on Covid-19 compatible symptoms was included in the composite primary endpoint.

The definition of Covid-19 compatible symptoms was based on guidance from the US Council for State and Territorial Epidemiologists **(Appendix)**.(12) Specifically, probable disease was defined as having cough, shortness of breath, or difficulty breathing, OR two or more of the following symptoms: fevers, chills, rigors, myalgia, headache, sore throat, new olfactory and taste disorders. Possible disease was defined as one or more COVID-19-compatible symptoms. Three blinded infectious diseases physicians independently adjudicated cases of symptomatic participants based on the above criteria.

Secondary outcomes included incidence of confirmed SARS-CoV-2 detection, incidence of possible Covid-19, and incidence of hospitalization, death, or other adverse events. Study medication adherence and side effects were all collected through weekly self-reported surveys.

### Randomization

Participants were sequentially randomized at the research pharmacies. Treatment assignments were concealed from investigators and participants. Blinded hydroxychloroquine or placebo (folic acid) was dispensed and shipped to participants by courier.

### Sample Size

The trial was designed anticipating a 10% event rate of Covid-19 in high-risk healthcare workers over 12 weeks. Using Log-rank testing with a 50% relative effect size to reduce new symptomatic infections, a two-sided alpha of 0.025 and 80% power, an estimated 1050 participants per arm were required. The trial was powered at a = 0.025 to account for the two treatment dosing regimens versus placebo comparisons.

### Statistical Methods

We compared the incidence of Covid-19–free survival using the Log-rank test and estimated hazard ratios using a Cox proportional hazards model. We compared secondary endpoints of proportions by Fisher’s exact test. We conducted analyses with SAS software version 9.4 (SAS Institute), according to the intention-to-treat principle. Participants were right-hand censored at time of last contact for those not completing 12 weeks of follow up. As prespecified, participants who developed Covid-19–compatible illness (i.e., primary endpoint) prior to initiating the study medicine were excluded from the primary analysis.

### Hydroxychloroquine drug concentrations

A prespecified subgroup analysis was performed to investigate whether hydroxychloroquine drug concentrations correlated with protection from Covid-19. Whole blood was self-collected from participants who consented using Neoteryx® volumetric absorbed microsampling kits (Neoteryx, Torrance, CA) at least four weeks after study medication initiation. Hydroxychloroquine concentrations were quantified similarly to methods previously published (**Appendix**).(13) The Wilcoxon rank-sum test compared trough concentrations between participants who developed Covid-19 and those who did not.

### Interim Analysis

An independent DSMB reviewed the data after 25% of participants had completed four weeks of follow up. Stopping guidelines were provided to the DSMB via a Lan-DeMets spending function analog of the O’Brien-Fleming boundaries for the primary outcome.

Before the first interim analysis on May 21, 2020, it became apparent that we would not meet our initial enrollment goal of 3150 participants (**Supplemental Figure S2**). At the first interim analysis, and without unblinding of treatment allocation, the principal investigator proposed to the data safety monitoring board stopping enrollment due to an inability to recruit participants, with continued follow up for those already enrolled. Enrollment was stopped on May 26, 2020, and outcomes data were collected through July 13, 2020.

## Results

Of 2271 persons screened, 1483 high-risk healthcare workers from the United States and Canada were enrolled with: 494 randomized to once-weekly hydroxychloroquine, 495 randomized to twice-weekly hydroxychloroquine, and 494 randomized to placebo (**Figure 1**). Participant demographics are provided in **Table 1**. The median age of participants was 41 years (interquartile range, 34 to 49), and 51% (760 of 1483) were women. Overall, 66% reported no chronic medical conditions (982 of 1483), while 14% (205 of 1483) reported hypertension and 10% asthma (150 of 1483). The primary location of work was the emergency department for 41% (607 of 1483), intensive care units for 18% (269 of 1483), operating rooms for 12% (178 of 1483), Covid-19 hospital wards for 10% (154 of 1483), and ambulance/first response teams for 8% (118 of 1483).

**Table 1.** Baseline Demographics.

| Characteristic | Placebo | Hydroxychloroquine<br>once weekly | Hydroxychloroquine<br>twice weekly |
| --- | --- | --- | --- |
| Number Randomized | 494 | 494 | 495 |
| Age in years, median (IQR) | 40 (34, 48) | 42 (35, 49) | 41 (35, 49) |
| Weight in kg, median (IQR) | 80 (68, 95) | 79 (67,93) | 82 (68, 95) |
| Female* –no. (%) | 241 (48.8%) | 261 (52.8%) | 258 (52.1%) |
| Ethnicity (all that apply) –no. (%) |  |  |  |
| White or Caucasian | 419 (84.8%) | 431 (87.2%) | 421 (85.1%) |
| Black or African | 10 (2.0%) | 5 (1.0%) | 5 (1.0%) |
| Asian | 29 (5.9%) | 23 (4.7%) | 23 (4.6%) |
| Native Hawaiian or Pacific Islander | 1 (0.2%) | 0 (0.0%) | 1 (0.2%) |
| Hispanic or Latino | 18 (3.6%) | 18 (3.6%) | 22 (4.4%) |
| Native American or Alaska Native | 8 (1.6%) | 4 (0.8%) | 7 (1.4%) |
| Middle Eastern | 4 (0.8%) | 6 (1.2%) | 5 (1.0%) |
| South Asian | 12 (2.4%) | 17 (3.4%) | 18 (3.6%) |
| Other | 4 (0.8%) | 3 (0.6%) | 1 (0.2%) |
| Current Smoker –no. (%) | 13 (2.6%) | 17 (3.4%) | 21 (4.2%) |
| Chronic Health<br>Conditions –no. (%) |  |  |  |
| High blood pressure | 60 (12.1%) | 79 (16.0%) | 66 (13.3%) |
| Asthma | 59 (11.9%) | 46 (9.3%) | 45 (9.1%) |
| None | 336 (68.0%) | 311 (63.0%) | 335 (67.7%) |
| **Covid-19 risk factors at screening** |  |  |  |
| Interacted with Covid19 patients when NOT wearing a<br>mask OR face shield? –no. (%) |  |  |  |
| Yes | 62 (12.6%) | 69 (14.1%) | 85 (17.2%) |
| No | 432 (87.4%) | 422 (85.9%) | 409 (82.8%) |
| Perform aerosol-generating procedures? –<br>no. (%) | 410 (83.0%) | 378 (77.0%) | 377 (76.3%) |
| Aerosol-generating procedures performed<br>per week –mean (SD) | 10 (31.4) | 9 (12.2) | 9 (12.5) |
| Setting of occupational<br>exposure –no. (%) |  |  |  |
| Emergency Department | 190 (38.5%) | 210 (42.5%) | 207 (40.8%) |
| Intensive Care Unit | 85 (17.2%) | 82 (16.6%) | 102 (20.6%) |
| Operating Room | 75 (15.2%) | 61 (12.3%) | 42 (8.5%) |
| COVID-19 ward | 56 (11.3%) | 51 (10.3%) | 47 (9.5%) |
| Ambulance | 45 (9.1%) | 40 (8.1%) | 33 (6.7%) |
| Congregate Care Setting | 20 (4.0%) | 19 (3.8%) | 27 (5.5%) |

\*No pregnant women enrolled, 30 women reported breastfeeding at baseline.

**Figure 1.**
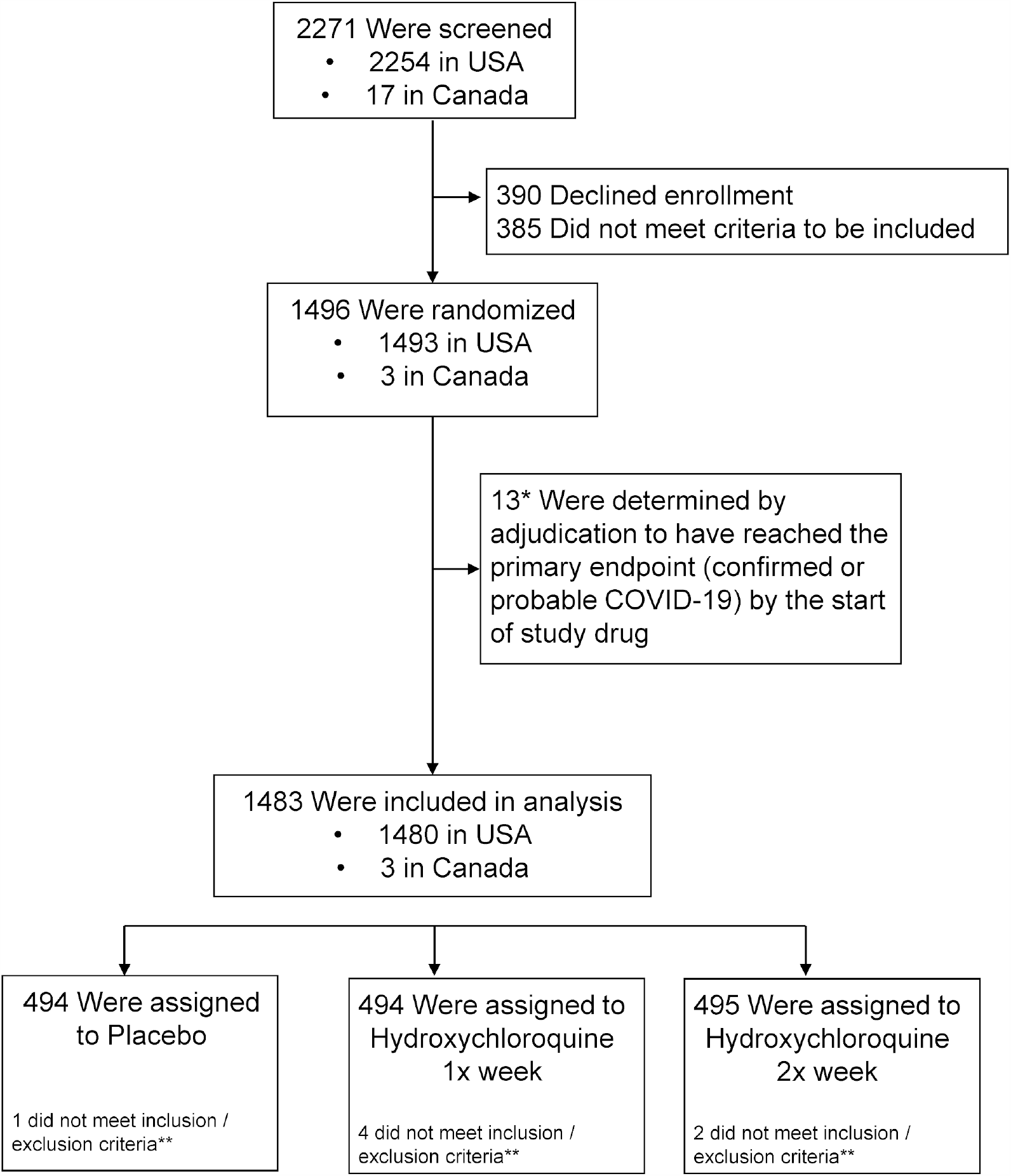
CONSORT Diagram.

Overall, 91% reported more than 14 hours of direct contact with patients per week (1346 of 1483), and 79% of participants reported routinely performing aerosol-generating procedures (1165 of 1483), with an average of nine procedures performed per week. For aerosol-generating procedures, 74% reported typically wearing an N95 respirator or powered air-purifying respirator (PAPR).

### Primary Outcome

The study accrued 311 person-years of follow up, and 97 participants (6.5%) developed Covid-19 (either PCR confirmed or symptomatically compatible illness) during the trial. Overall, confirmed or probable Covid-19–compatible illness occurred in 29 (5.9%) receiving once-weekly hydroxychloroquine, 29 (5.9%) receiving twice-weekly hydroxychloroquine, and 39 (7.9%) receiving placebo. The corresponding incidence of Covid-19 or compatible illness was 0.27 and 0.28 events per person-year for those taking hydroxychloroquine once or twice weekly, respectively, as compared to 0.38 events per person-year in those receiving placebo (**Table 2**). Compared to placebo, the hazard ratios for Covid-19 or compatible illness were 0.72 (95%CI 0.44 to 1.16; P=0.18) with once-weekly and 0.74 (95%CI, 0.46 to 1.19; P=0.22) with twice-weekly hydroxychloroquine respectively (**Figure 2**). When hydroxychloroquine arms were combined, the hazard ratio for Covid-19 or compatible illness was 0.73 (95% CI 0.48 to 1.09; P=0.12) compared with placebo.

**Table 2.** Incidence of Covid-19 with hydroxychloroquine as pre-exposure prophylaxis.

| **Outcome** | **Placebo** |  | **Hydroxychloroquine once weekly** |  |  | **Hydroxychloroquine twice weekly** |  |  |
| --- | --- | --- | --- | --- | --- | --- | --- | --- |
|  | No. of infections (%) | Event rate per person-yr (95% CI) | No. of infections (%) | Event rate per person-yr (95% CI) | Hazard Ratio (95% CI) | No. of infections (%) | Event rate per person-yr (95% CI) | Hazard Ratio (95% CI) |
| PCR positive or probable Covid-19 | 39 (7.9%) | 0.38 (0.26 to 0.50) | 29 (5.9%) | 0.27 (0.17 to 0.37) | 0.72 (0.44 to 1.16) | 29 (5.9%) | 0.28 (0.18 to 0.38) | 0.74 (0.46 to 1.19) |
| PCR confirmed Covid-19 | 6 (1.2%) | 0.06 (0.01 to 0.10) | 4 (0.8%) | 0.04 (0.00 to 0.07) | 0.65 (0.18 to 2.32) | 7 (1.4%) | 0.07 (0.02 to 0.12) | 1.18 (0.40 to 3.51) |
| Covid-19 compatible with symptoms | 38 (7.7%) | 0.38 (0.26 to 0.49) | 29 (5.9%) | 0.28 (0.18 to 0.38) | 0.73 (0.45 to 1.19) | 28 (5.7%) | 0.28 (0.17 to 0.38) | 0.74 (0.45 to 1.20) |

**Figure 2:**
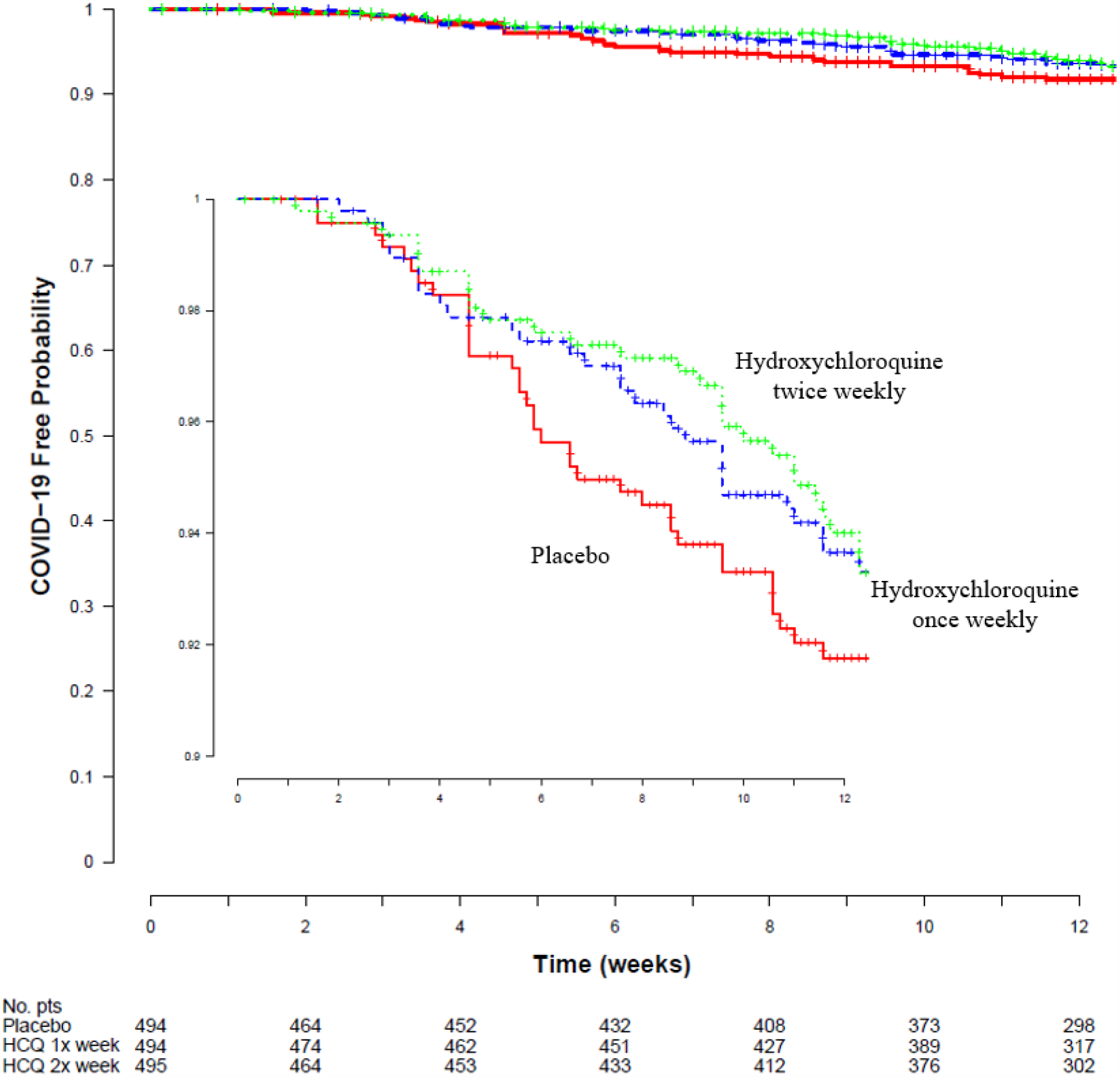
Kaplan Meier Estimates of Time to Covid-19 Compatible illness. The probability of SARS CoV-2 infection over time is shown for the three study groups. The hazard ratio for twice weekly hydroxychloroquine prophylaxis was 0.72 (95%CI 0.44 to 1.16; p= 0.18) and for once weekly hydroxychloroquine was 0.74 (95%CI 0.46 to 1.19; p=0.22) as compared with placebo. The inset graph shows more detail.

Of the 97 with Covid-19 (PCR confirmed or symptomatically compatible illness, 17 were PCR positive (18%), 42 (43%) had no PCR testing during their illness, and 38 (39%) had a negative PCR test during illness (**Appendix**). Of the 38 with a negative PCR, 30 were collected within 4 days prior to symptom onset, and 8 collected within 11 days after symptom onset. The hazard ratio for PCR-confirmed Covid-19 was 0.65 (95% CI 0.18 to 2.32, P=0.51) for once-weekly hydroxychloroquine, and 1.18 (95% CI 0.40 to 3.51, P=0.77) for twice-weekly hydroxychloroquine. (**Table 2**).

### Medication Adherence and Side Effects

Self-reported adherence to study medicine was not significantly different by treatment group (**Figure S9, Appendix**). Of those who reported full adherence at ≥80% of surveys, Covid-19 occurred in 8.5% (28/331) of participants assigned to placebo, 5.7% (20/351) of participants assigned to once-weekly hydroxychloroquine (Hazard Ratio 0.66, 95% CI 0.37 to 1.17; P=0.16), and 5.7% (18/316) of participants assigned to twice-weekly hydroxychloroquine (Hazard Ratio 0.68, 95% CI 0.37 to 1.22; P=0.19).

Side effects were reported in 21% of participants assigned to placebo (100 of 469) (**Supplemental Table 4**), 31% in the once-weekly hydroxychloroquine group (148 of 473; P<0.001), and 36% in the twice-weekly hydroxychloroquine group (168 of 463; P<0.001). The most common side effect was stomach upset and nausea (placebo 12.2%, hydroxychloroquine once-weekly 17.5%, and hydroxychloroquine twice-weekly 19.4%), followed by gastrointestinal disturbance and diarrhea (placebo 7.5%, hydroxychloroquine once-weekly 12.9%, and hydroxychloroquine twice-weekly 17.1%).

### Other Secondary Outcomes

Twenty hospitalizations occurred during the study: nine in the placebo arm, three in the hydroxychloroquine once-weekly arm, and eight in the hydroxychloroquine twice-weekly arm. Reasons for hospitalization are summarized in the **Appendix**. Two hospitalizations were related to Covid-19 (1 placebo, 1 twice-weekly group). One person in the placebo group was hospitalized twice for new atrial fibrillation, and one person in the hydroxychloroquine twice-weekly arm was hospitalized for syncope and new supraventricular tachycardia – a possible hydroxychloroquine-related serious adverse event (SAE)). No intensive care unit stays, or deaths occurred in this study.

In prespecified subgroup analyses, there were no significant differences in treatment efficacy (**Appendix**).

### Hydroxychloroquine drug concentrations

Hydroxychloroquine concentrations were measured in dried whole blood from 180 participants in the hydroxychloroquine groups of whom 18 were confirmed or probable Covid-19 and 6 were considered possible Covid-19 **(Supplemental Table 7)**. Hydroxychloroquine was detectable in all samples analyzed. Median (Interquartile range (IQR)) concentrations were higher in the twice-weekly dosing group [200 ng/mL (IQR, 159-258)] compared to the once-weekly dosing group [98 ng/mL (IQR, 82-120; P<0.0001)]. Median concentrations did not differ between Covid-19 confirmed, probable or possible cases [154 (IQR, 119-231) ng/mL] compared with participants without Covid-19, [133 (IQR, 93-198) ng/mL, (P=0.08)] (**Figure 3)**.

**Figure 3:**
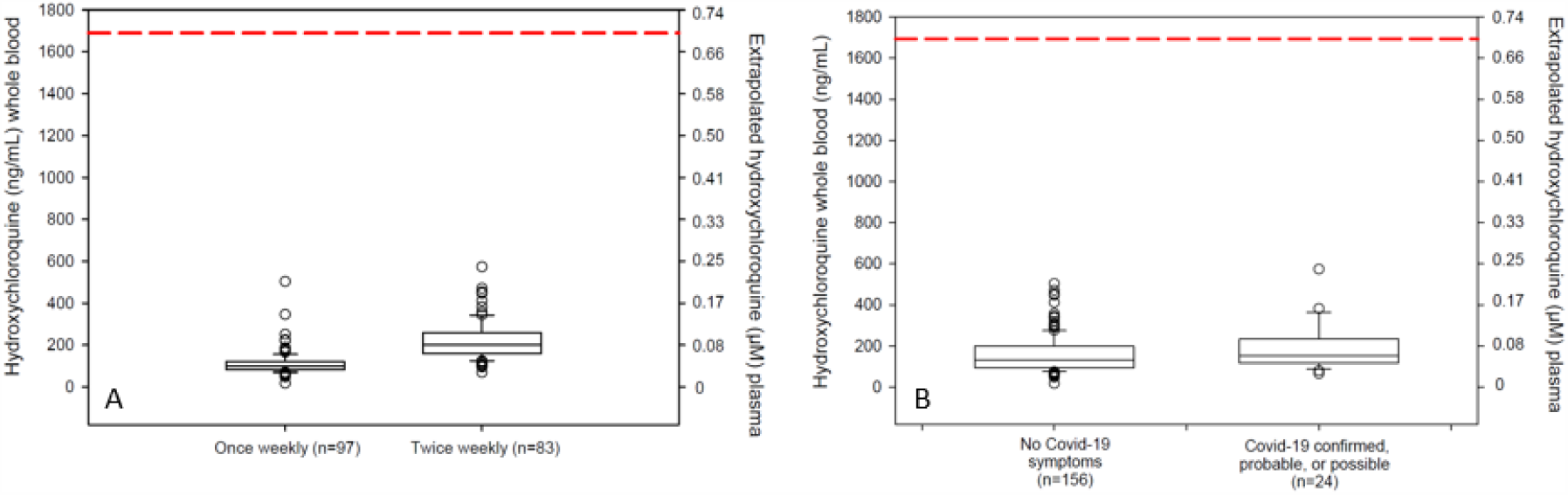
Hydroxychloroquine drug concentrations. Right-side axes indicate extrapolated plasma concentrations assuming blood to plasma ratio of 7.2 and hydroxychloroquine molecular weight of 336 g/mol.(16) Panel A shows trough drug concentrations in participants taking once weekly compared with twice weekly hydroxychloroquine. All participants had detectable hydroxychloroquine in whole blood samples. Panel B depicts drug concentrations in participants from both hydroxychloroquine arms with and without Covid-19 compatible illness. Participants with Covid-19 compatible illness had median concentrations of 154 ng/mL compared with 133 ng/mL among those without symptomatic Covid-19 compatible illness (P=0.08). Red dashed line indicates extrapolated EC50 target assuming blood to plasma ratio of 7.2, target EC50 of 0.7μm = 235 ng/mL plasma = 1690 ng/mL whole blood.(5)

## Discussion

In this randomized, double-blind, placebo-controlled trial evaluating hydroxychloroquine as pre-exposure prophylaxis for Covid-19 in high-risk healthcare workers, we found no statistically significant reduction in Covid-19 incidence in those receiving 400mg weekly or twice weekly hydroxychloroquine when compared with placebo. Reasons for no effect observed may be due to hydroxychloroquine levels being too low, or because hydroxychloroquine is ineffective against Covid-19 in vivo.(14)

Nonetheless, we observed no difference in hydroxychloroquine concentrations between those who reported Covid-19 symptomatically compatible illness and those who did not, in a subsample of trial participants. Similarly, an animal model of macaques showed that hydroxychloroquine offered no protection against SARS-CoV-2 acquisition when given as pre-exposure prophylaxis.(14) While there are no validated therapeutic target concentrations of hydroxychloroquine for protection against Covid-19, we chose dosing regimens predicted to achieve plasma concentrations above the *in vitro* EC50.(15) Assuming blood concentrations are seven-fold higher than plasma, (16) no participants had plasma troughs higher than reported *in vitro* EC50. Plasma concentrations of 235 ng/mL (∼0.7μM) would extrapolate to a whole blood target >1600 ng/mL, significantly higher than troughs achieved in our study. The discrepancy between our simulated and observed concentrations is consistent with a recent analysis,(17) which suggested that due to sequestering of drug in whole blood leukocytes and thrombocytes not adequately removed during processing, the pharmacokinetic parameters upon which we based our simulations may have overestimated plasma concentrations.(18) This finding is likely applicable to all hydroxychloroquine trials. Notably, our whole blood troughs suggest that even with daily dosing, extrapolated plasma trough concentrations above EC50 are unlikely. Ongoing trials investigating the efficacy of daily dosing should consider obtaining plasma levels to further decipher whether daily dosing is adequate, and in the context of appropriate dosing, if hydroxychloroquine is effective at preventing SARS CoV-2 infection. Our results suggest that prophylaxis with hydroxychloroquine 400mg weekly is ineffective, and recommendations for prophylactic use, such as those for healthcare workers in India, should be reconsidered.

When justifying widespread implementation of a prophylactic intervention, it is paramount to consider and predefine a required minimum efficacy. Unfortunately, there are no evidence-based guidelines on Covid-19 prophylaxis, given its rapidly emerging and ongoing nature. When designing this study in March 2020, we used the human immunodeficiency virus (HIV) as a model for pre-exposure prophylaxis to hypothesize what would constitute a successful prophylactic intervention warranting widespread implementation. Daily oral pre-exposure prophylaxis in HIV has previously been associated with a 92% relative reduction in HIV acquisition among participants with detectable study medication levels.(19) Together with the Food and Drug Administration (FDA)’s suggestion that a minimum efficacy of 50% was required for a Covid-19 vaccine to be approved, we hypothesized that a 50% relative risk reduction in confirmed or probable Covid-19 would be clinically meaningful and powered the study design as such. Our estimates of incidence of Covid-19 (confirmed or symptomatic) will be valuable for future studies of chemoprophylaxis and vaccine trials.

Enrolling participants was a challenge. We enrolled 84% of all participants (1250 of 1483) in the first two weeks of the trial. During April 21-24, 2020, a series of small or retrospective studies highlighted safety concerns of hydroxychloroquine,(20, 21) which resulted in a warning from FDA regarding arrhythmias and QT prolongation.(22) Thereafter, our enrollment precipitously declined. An additional study in May, which is now retracted,(23) further discouraged enrollment. Enrollment was stopped on May 26, 2020, due to futility in ongoing participant recruitment. Enrollment in all other North American randomized clinical trials of hydroxychloroquine was also significantly impeded (Dee Dee Wang, personal communications). As a result of premature enrollment termination and inadequate power, it is difficult to estimate the potential societal benefit, if any, in widespread implementation. Based on the demonstrated absolute risk reduction of 0.11 events per person-years, nine high-risk healthcare workers would need to receive prophylaxis for one year to prevent one Covid-19 case. The potential benefit would be less in healthcare workers at lower risk and in the general population at large due to fewer exposure events; however, the potential benefit would vary based on local prevalence of Covid-19.

The major limitation of this trial relates to the inherent challenges with PCR testing that have been well described-both the lack of U.S. availability and moderate reported sensitivity early in illness. The false-negative rate of PCR testing has been reported to be 38% (Range 18-65%) on the first day of symptoms, gradually decreasing thereafter.(24) In our study, 39% (38/97) had Covid-19-compatible symptoms with a negative PCR test; however 30 of those PCR tests were performed before symptoms began, when false negatives can be expected.(24) To address this, we included healthcare workers with symptomatic Covid-19–compatible illness despite negative PCR, but separately reported this group. Further supporting this decision, Covid-19–compatible symptoms warrant self-isolation from work for 14 days for healthcare providers and reporting to occupational health, per CDC guidelines, even if PCR testing is negative.(25) However, it is unknown what proportion of persons with symptomatically compatible disease truly have SARS CoV-2 infection, which remains a shared limitation to all outpatient Covid-19 trials in the absence of a diagnostic test with improved sensitivity.

Hypothetically, if reported symptoms were due to another respiratory illness, such as influenza, they should have been evenly distributed between groups due to randomization. If one compares only PCR-confirmed disease, there was no statistical difference between groups. Secondly, our trial was limited by weekly self-report of outcomes, subject to recall bias. As mentioned previously, insufficient dosing of hydroxychloroquine remains a limitation of this study. Finally, our trial was left underpowered due to impediments to participant recruitment. Nevertheless, the effect size estimates derived from our data will inform current policy and aid in the design of future clinical trials.

An effective means of prophylaxis for high-risk healthcare workers remains a critical need in the context of a growing and relentless pandemic. The COVID PREP study evaluated the effectiveness of once-weekly and twice-weekly hydroxychloroquine to prevent Covid-19 in high-risk healthcare workers across the United States and Canada. There was no statistically significant reduction in the incidence of Covid-19 in our trial. However, investigation into more frequent dosing may be warranted. Prior to embarking on further clinical trials, and for current studies to complete enrollment, the perception of equipoise in the medical community and the public will need to change dramatically.

## Data Availability

Data will be available 1 month after publication in a peer-reviewed journal.

https://covidprep.umn.edu/

## Disclosure

The funders did not contribute to the design, collection, management, analysis, interpretation of data, writing of the report, nor the decision to submit the report for publication.

## Acknowledgements

We thank the healthcare workers around North America who volunteered to participate in this trial in order to obtain knowledge for society. We thank the thoughtful service of DSMB members: Drs. Mark Seidner, Lynn Matthews, Jeff Klausner, Bozena Morawski, and Tom Chiller. We thank institutional support from Drs. Jakub Tolar, Peter Igarashi, Brad Benson, and Tim Schacker.

This work was supported by Jan and David Baszucki, Steve Kirsch, the Rainwater Charitable Foundation, the Alliance of Minnesota Chinese Organizations, the Minnesota Chinese Chamber of Commerce, and the University of Minnesota Foundation. Drs. Melanie Nicol, Radha Rajasingham, and Matthew Pullen are supported by the National Institute of Allergy and Infectious Disease (K08AI134262, K23AI138851, T32AI055433). Sarah Lofgren is supported by the National Institute of Mental Health (K23MH121220). Caleb Skipper is supported by a combined Fogarty International Center/National Institute of Neurological Disorders and Stroke grant (D43TW009345). Katelyn Pastick and Elizabeth Okafor were supported through the Doris Duke Charitable Foundation through a grant supporting the Doris Duke International Clinical Research Fellows Program at the University of Minnesota. Margaret Axelrod is supported by NIH T32GM007347 and F30CA236157. Drs. Lee and McDonald receive research salary support from the Fonds de recherche du Québec – Santé. In Manitoba, research support was received from the Manitoba Medical Service Foundation and Research Manitoba. Rising Pharmaceuticals in the U.S. provided a donation of the hydroxychloroquine tablets. The REDCap software was supported by the National Institutes of Health’s National Center for Advancing Translational Sciences, grant UL1TR002494. The funders had no role in trial design, trial implementation, writing of the manuscript, or the decision to submit it for publication.

**<u>COVID PREP team members</u>** (listed alphabetically)

Mahsa Abassi, DO. University of Minnesota, Minneapolis, MN.

Andrew Balster, MD. Oregon Health & Science University, Portland, OR.

Lindsey B. Collins, BSc. University of Minnesota, Minneapolis, MN.

Glen Drobot, MD. University of Manitoba, Winnipeg, Manitoba.

Douglas S. Krakower, MD. Beth Israel Deaconess Medical Center, Boston, MA.

Sylvain A. Lother, MD. University of Manitoba, Winnipeg, Manitoba.

Dylan S. MacKay, PhD. University of Manitoba, Winnipeg, Manitoba.

Cameron Meyer-Mueller, BA. University of Minnesota, Minneapolis, MN.

Stephen Selinsky, MD. University of Minnesota, Minneapolis, MN.

Dayna Solvason. The George and Fay Yee Centre for Healthcare Innovation, Winnipeg, Manitoba.

Ryan Zarychanski, MD, MSc. University of Manitoba, Winnipeg, Manitoba.

Rebecca Zash, MD. Beth Israel Deaconess Medical Center, Boston, MA

## Collaborators

Drug assay development and performance: James Fisher

Concept advisors: Archana Bhaskaran

Logistical Support: Kristen Moran, Alek Lefevbre, Izabella Supel, Carmen Tse, Hongru Ren,

Fiona Vickers, Jason Zou: Pharmacy Support

Darlette Luke, PRISM Research Inc., Halyna Ferens, Beata Kozak

## Authorship Contributions

RR and DRB conceived of the trial. RR wrote the clinical protocol with the assistance of SML, CPS, DRB, MRN, JB,BR, PL and IM, and statistical input from ASB, NWE, and KHH. LJM collaborated and adapted the study for Canadian sites with input from DSM, DS and TCL. KHH, ASB, and NWE conducted the statistical analyses, with the analysis being guaranteed by KHH. ASB developed the REDCap database with help from MFP, KAP, SML, and CPS. ASB maintained the database. TCL and EGM adapted the database in Canada. MLA, CPS, AAN, MFP, KAP, ECO, DAW, LJM, and RR did participant follow-up. Advertising and outreach were done by SD, DRB, RR, SML, CPS, MFP, ASB, AAN, ECO, PL, DAW, LJM, SAL, DSM, GD, and RZ. RR, CPS and SML did case adjudication. Hydroxychloroquine drug levels sub study was conceived by MRN, RR, DK, RZ, and levels were analyzed by MRN. RR wrote the first draft of the manuscript and is the overall study guarantor with help from SML, DRB, CPS, MFP, KAP, ECO, AAN, and DAW. All authors reviewed and revised, and approved the final version of the manuscript. The FDA Investigational New Drug sponsor is RR.

Steve Kirsch, David Baszucki and Jan Ellison Baszucki, the Rainwater Charitable Foundation, the Alliance of Minnesota Chinese Organizations, the Minnesota Chinese Chamber of Commerce, and the University of Minnesota provided funding but did not have a role in protocol development or monitoring. Rising Pharmaceuticals provided the hydroxychloroquine tablets.

## Notes

### Competing Interest Statement

The authors have declared no competing interest.

### Clinical Trial

NCT04328467

### Author Declarations

This trial was approved by the University of Minnesota Institutional Review Board, conducted under FDA Investigational New Drug (149252), and overseen by an independent DSMB. In Canada, the trial was authorized without objection by Health Canada (control number 238396), with ethics approval obtained from the University of Manitoba.

